# Estimating ascending aortic diameter from the electrocardiogram

**DOI:** 10.1101/2025.08.19.25333786

**Authors:** Zachariah S. Demarais, Jeffrey E. Olgin, James P. Pirruccello

## Abstract

In an analysis of 69,173 UK Biobank participants, we paired MRI-based measurements of the ascending aortic diameter with ECG signal. We trained a 1D convolutional neural network (ECGAI-TAA) to consume the 10-second 500Hz 12-lead signal and to emit an estimate of the ascending aortic diameter. We assessed model performance in an internal test set of 5,191 participants. The resulting model explained 31% of the variance in aortic diameter, and it couldn’t be fully explained by clinical factors such as age, sex, height, weight, pulse rate, blood pressure, or left ventricular mass. Evaluating a clinically relevant diameter threshold (4.0cm, representing dilation), 2.5% of the population had a dilated ascending aorta; when comparing that same proportion of the population (individuals in the top 2.5% of the deep learning model score) to the remaining participants, we found a nearly 16-fold odds ratio for aortic dilation.

Using a variational autoencoder-based visualization, we hypothesized that a lateral-superior axis shift may underlie the electrical changes being detected by the model. An important limitation is that these findings represent a physiological observation, not an externally validated risk score. In conclusion, the ECGAI-TAA deep learning model demonstrates that ascending aortic diameter can be, in part, estimated from the 12-lead ECG.

## Main text

The electrocardiogram (ECG) is commonly used for assessment of arrhythmia and myocardial dysfunction or injury. Recent discoveries have shown that deep learning models of ECG can also detect abnormalities that are not strictly myocardial, such as valvular heart disease^1^. Here, we sought to understand whether ascending aortic diameter can be estimated from the ECG.

This research has been conducted using the UK Biobank Resource under Application Number #41664, with UCSF IRB approval #22-37715. After excluding any participants with resting heart rate below 20 or above 100, we analyzed 69,173 UK Biobank participants with 12-lead ECG and ascending aortic diameter measurements from magnetic resonance imaging (MRI)^2^. With PyTorch v2.7.1^3^, a 1-dimensional ResNet-like ECG deep learning model for the thoracic ascending aorta (“ECGAI-TAA”) was trained. The input was composed of 10-second 12-lead 500Hz ECG signals. The model had a stem with kernel size 15; three residual stages with group normalization and downsampling between blocks; dilations in alternating blocks; a residual multi-head attention block; and finally global average pooling, dropout, and a linear layer projecting onto ascending aortic diameter and heart rate. The training loss function was the mean squared error for normalized aortic diameter and heart rate; for validation, only aortic diameter was considered. Data from 62,255 participants (90%) were used for model training, from 1,727 (2.5%) for validation, and from 5,191 (7.5%) for a final internal test set. For data augmentation during training, noise was added to each lead. The ECG signal was left in its millivolt (mV) scale without preprocessing, and the ascending aortic diameter was measured in units of centimeters (cm). Training was conducted with a batch size of 64. After a linear warmup of 2,000 minibatches, a flat learning rate of 1e-3 was used for 30,000 minibatches. Validation was assessed every 500 minibatches; the model weights with the lowest validation loss were saved. Inference was then run to generate ECGAI-TAA scores for all test-set participants (**Figure Panel A**).

An *R* v4.5.0 linear model was used to assess the relationship between the ECGAI-TAA score (representing the estimated ascending aortic diameter in cm) and the MRI-based ascending aortic diameter measurements. This yielded an estimate of a diameter of 0.97cm per 1.0cm of the ECGAI-TAA score, P=3.2E-423, R^2^=0.31 (explaining 31% of the variance in diameter). For context, a linear model accounting for age and sex alone explained 17.9% (P=9.3E-223) of the variance in ascending aortic diameter, which increased to 32.7% (P=6.5E-446) after incorporating the ECGAI-TAA score. A model accounting for age, sex, height, weight, heart rate at the time of MRI, systolic blood pressure, and diastolic blood pressure (N=4,539 complete cases) explained 28.0% (P=9.7E-319) of the variance in ascending aortic diameter, which increased to 37.2% (P=1.7E-452) after incorporating the ECGAI-TAA score. When additionally accounting for left ventricular (LV) mass, the model without ECG accounted for 33.0% of the variance (P=4.4E-385) and 39.1% with ECG (P=4.7E-477).

130 of the 5,191 test-set participants (2.5%) had ascending aortic dilation (diameter ≥ 4cm). On average, these participants had an ECGAI-TAA score that was 1.42 standard deviations greater (95% confidence interval [CI] 1.25-1.59) than that of all other participants. For each standard deviation greater ECGAI-TAA score, the odds ratio (OR) for aortic dilation was 3.6 (95% CI 3.0- 4.3). 31 participants in the top 2.5% of the ECGAI-TAA score also had an aortic diameter ≥ 4.0cm, yielding an OR of 15.7 (95% CI 10.0-24.6) for identifying ascending aortic dilation (**Figure Panel B**).

To develop a qualitative hypothesis around the ECG features that might be relevant for ascending aortic diameter estimation, we trained a variational autoencoder (VAE) using the 12- lead median waveforms from the same participants. The VAE was modeled after Representation Learning for Genetic Discovery on Low-Dimensional Embeddings [REGLE]^4^ and had a latent dimension of 128, plus one injected dimension for aortic diameter. After training for 350,000 minibatches, the trained model was applied to data from one individual in the test set.

The latent vector was fixed, and the aortic diameter parameter was varied from 2.3-4.1cm. The generated output is shown in **Figure Panel C**.

To our knowledge, this work represents the first observation that ascending aortic diameter can be estimated from the ECG. The ascending aorta acts as a conduit for blood and an elastic buffering system for cardiac energy, but by bulk mass it is largely composed of electrically inert material. Consequently, we expect that these observations represent electrically detectable epiphenomena rather than a direct readout of aortic diameter. The sensitivity analysis that accounted for LV mass suggested that the ECGAI-TAA model is not simply inferring LV mass as a proxy for aortic diameter. Using the VAE for qualitative hypothesis generation, we noted that the salient phenomenon being captured by the EKG models may be one of lateral and superior axis displacement (especially given the greater increase in lead aVL QRS voltage compared to lead I). If mechanical, this may represent a left-superior displacement of the cardiac long-axis, plausibly due to a pivot around the great vessels. It is important to emphasize that the VAE does not represent a causal model. Future efforts will be required to clarify the extent to which a mechanical—rather than purely electrophysiologic—effect is being observed. It will be of interest to determine whether ECG models will externally generalize and yield improvement when incorporated into models such as the AORTA Score^5^ and AORTA Gene^2^ to facilitate screening for sporadic ascending aortic aneurysm.

In conclusion, the ECGAI-TAA deep learning model demonstrates that ascending aortic diameter can be estimated from the 12-lead ECG.

**Figure 1.**
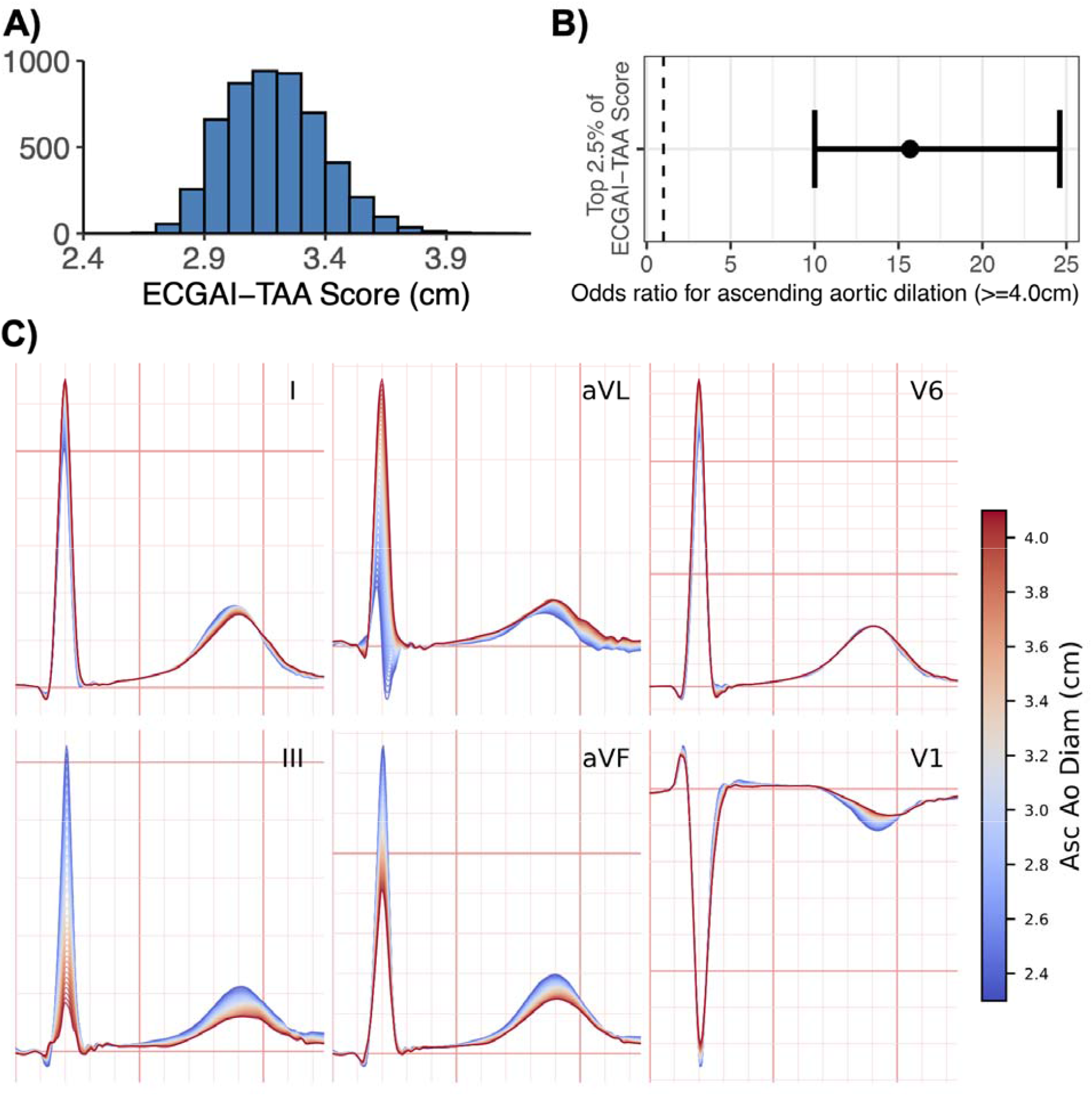
**Panel A**: Distribution of the ECGAI-TAA score estimates (with units of 1 cm) in the 5,191 test set participants. **Panel B**: For the top 2.5% of test set participants by ECGAI-TAA score, odds ratio and 95% confidence interval for presence of ascending aortic dilation (diameter ≥ 4.0cm). **Panel C**: Selected reconstructed QRS complexes from a variational autoencoder conditioned on ascending aortic diameter. Inference was repeatedly run on data from one ECG, replacing the true aortic diameter with values from 2.3-4.1cm; lines are colored based on these diameters. Standard ECG gridlines are shown (40ms per step along the x-axis, 0.1mV per step along the y-axis). For each lead, the y-axis scale was allowed to vary to emphasize detail. Derived ECG signals reproduced by kind permission of UK Biobank ©.

## Data Availability

UK Biobank data are available to researchers with UK Biobank approval.

## Acknowledgments

J.P.P. reports funding from the NIH’s NHLBI (R01HL178603 and K08HL159346).

## Notes

### Author Declarations

IRB of University of California San Francisco gave ethical approval for this work (#22-37715)

